# Sample average treatment effect on the treated analysis using counterfactual explanation identifies BMT and SARS-CoV-2 vaccination as protective risk factors associated with COVID-19 severity and survival in patients with multiple myeloma

**DOI:** 10.1101/2022.12.07.22283208

**Authors:** Amit Kumar Mitra, Ujjal Kumar Mukherjee, Suman Mazumder, Vithal Madhira, Timothy Bergquist, Yu Raymond Shao, Feifan Liu, Qianqian Song, Jing Su, Shaji Kumar, Benjamin A. Bates, Noha Sharafeldin, Umit Topaloglu, the National COVID Cohort Collaborative Consortium

## Abstract

Patients with multiple myeloma (MM), an age-dependent neoplasm of antibody-producing plasma cells, have compromised immune systems and might be at increased risk for severe COVID-19 outcomes. This study characterizes risk factors associated with clinical indicators of COVID-19 severity and all-cause mortality in myeloma patients utilizing NCATS’ National COVID Cohort Collaborative (N3C) database. The N3C consortium is a large, centralized data resource representing the largest multi-center cohort of COVID-19 cases and controls nationwide (>16 million total patients, and >6 million confirmed COVID-19+ cases to date).

Our cohort included myeloma patients (both inpatients and outpatients) within the N3C consortium who have been diagnosed with COVID-19 based on positive PCR or antigen tests or ICD-10-CM diagnosis code. The outcomes of interest include all-cause mortality (including discharge to hospice) during the index encounter and clinical indicators of severity (i.e., hospitalization/emergency department/ED visit, use of mechanical ventilation, or extracorporeal membrane oxygenation (ECMO)). Finally, causal inference analysis was performed using the propensity score matching (PSM) method.

As of 05/16/2022, the N3C consortium included 1,061,748 cancer patients, out of which 26,064 were MM patients (8,588 were COVID-19 positive). The mean age at COVID-19 diagnosis was 65.89 years, 46.8% were females, and 20.2% were of black race. 4.47% of patients died within 30 days of COVID-19 hospitalization. Overall, the survival probability was 90.7% across the course of the study. Multivariate logistic regression analysis showed histories of pulmonary and renal disease, dexamethasone, proteasome inhibitor/PI, immunomodulatory/IMiD therapies, and severe Charlson Comorbidity Index/CCI were significantly associated with higher risks of severe COVID-19 outcomes. Protective associations were observed with blood-or-marrow transplant/BMT and COVID-19 vaccination. Further, multivariate cox proportional hazard analysis showed that high and moderate CCI levels, International Staging System (ISS) moderate or severe stage, and PI therapy were associated with worse survival, while BMT and COVID-19 vaccination were associated with lower risk of death. Finally, matched sample average treatment effect on the treated (SATT) confirmed the causal effect of BMT and vaccination status as top protective factors associated with COVID-19 risk among US patients suffering from multiple myeloma.

To the best of our knowledge, this is the largest nationwide study on myeloma patients with COVID-19.

## INTRODUCTION

The coronavirus disease 19 (COVID-19) caused by Severe Acute Respiratory Syndrome Coronavirus 2 (SARS-CoV-2) has resulted in unprecedented consequences across the world in terms of mortality and quality of life^1^. Declared a pandemic by the WHO on March 11^th,^ 2020, COVID-19 has accounted for >1% of deaths globally in more than 180 countries, with several notable rapid surges (pandemic waves) across the world (https://covid19.who.int/) and multiple variant strains, notably B.1.1.7 (Alpha), B.1.351 (Beta), B.1.1.28 (P.1, Gamma) and B.1.617.2 (Delta) (https://www.cdc.gov/coronavirus/2019-ncov/variants/variant-classifications.html). The highly transmissible Omicron (B.1.1.529) variant that emerged in late 2021 spread in >75 countries and posed another serious threat to the already-dismal circumstances. Furthermore, multiple mutations in strain sublineages are a serious concern owing to their ability to surpass immunity (antibody evasion) and the degree of infectivity ^2^.

Cancer still remains one of the major causes of death worldwide, with a rapid increase in incidence, prevalence, and mortality over the recent decades (https://seer.cancer.gov/about/). Recent studies have shown that vulnerable cancer patients infected with COVID-19 present with more severe complications compared to healthy people living in the community^3^. Furthermore, several previous studies, including ours, have reported that the risk of death is also significantly higher in cancer patients^4^. Therefore, COVID-19-related deaths in cancer patients are highly challenging, more so because of the competing and unknown risks associated with active oncologic treatment as well as with delivering patient care.

Multiple myeloma (MM) is the second-most common hematopoietic malignancy in the United States^5^. MM is an age-dependent plasma cell neoplasm characterized by clonal expansion of malignant antibody-producing post-germinal-center B cell-derived plasma cells within the bone marrow^5^. Therefore, patients with hematological malignancies, particularly multiple myeloma, have compromised immune systems due to multiple factors, including comorbidities associated with the mean age of diagnosis at ∼65yo, loss of functional immunoglobulins, low CD4+T-cell count, suppression of normal B-cell development, as well as immunosuppression through immunomodulatory drugs/IMiDs^6^. This may increase the risk of severe SARS-CoV-2/COVID-19 infection and post-acute sequelae of SARS-CoV-2/PASC/long-COVID. Moreover, myeloma patients also present with a substantial multifactorial burden of cardiovascular disease, renal impairment, lymphopenia, neutropenia, and increased risk for venous thromboembolism/VTE that may be aggravated by pre-existing conditions, disease complications, and drug toxicities which are reported as risk factors among COVID-19 patients with a potentially fatal outcome^7,8^. In fact, an earlier study showed that myeloma patients experience 34% higher inpatient mortality due to COVID-19^9^. Although there are a handful of studies investigating how the incidence of COVID-19, its treatment and the interaction between COVID-19 and anti-myeloma therapies affect outcomes^9,10^there is a significant lack of studies that include substantially large datasets (>10000 myeloma patients).

In this study, we aim to expand the previous findings on the risk factors associated with COVID-19 severity and death and the impact of anti-myeloma therapy using a very large, naturally-representative cohort of cancer patients available through the National COVID Cohort Collaborative (N3C) initiative. The NCATS’ National COVID Cohort Collaborative/N3C is a centralized data resource representing the largest multi-center cohort of COVID-19 cases and controls nationwide^11^.

The NCATS’s N3C is the largest cancer cohort registry of COVID-19 tested patients nationwide that includes Electronic Health Record (EHR) data with at least one clinical encounter after January 1st, 2020^12^. As of July 31st, 2022, N3C houses centralized data on 14,778,237 patients from 73 contributing sites. This included 5,815,680 patients who tested positive for COVID-19. Our cohort study includes 26,064 myeloma patients, out of which 8,588 were confirmed COVID-19-positive. We used this large national-level clinical registry of myeloma patients with COVID-19 to identify predisposing and treatment-related factors associated with severity and all-cause mortality within our cohort.

## METHODS

### Study cohort

Our N3C myeloma cohort included patients (both inpatients and outpatients) from contributing sites who have been diagnosed with COVID-19 between January 1st, 2020, till our cut-off date May 16^th,^ 2022, 2022 (N3C release v76). All myeloma patients without COVID-19 encountered during this time period at the contributing sites were also included initially to build the overall myeloma cohort. Historical patient data from January 1st, 2018, were included for each patient from the same health system, wherever available.

### Indicator variables

The N3C clinical data set is a limited dataset that includes protected health information that may include dates of service and patient ZIP code. Details regarding data quality and harmonization checks, cohort definitions, and Malignant Neoplastic Disease standard (SNOMED) concept codes used for primary cancer diagnosis have been published earlier. Briefly, Cancer patients within the N3C registry were identified using the SNOMED Code 3633460000 by the Observational Health Data Sciences and the Informatics Atlas tool. For COVID-19 status, we used N3C positive phenotyping guidelines based on concept definitions and logic provided in **Supplementary Table 1**. For the purpose of this study, we limited our analysis to 30 days before the COVID-19 diagnosis to 30 days after the start of the index encounter. Further, we used available data to calculate indicator variables on the Charlson Comorbidity Index (CCI) adjusted for cancer diagnosis, primary cancer diagnosis, and cancer therapies.

### Myeloma diagnosis

International Staging System (ISS) for Multiple Myeloma stage was calculated using the revised guidelines provided by the International Myeloma Foundation (https://www.myeloma.org/international-staging-system-iss-reivised-iss-r-iss) as Stage 1: Alb ≥3.5, B2M< 3.5; Stage 2: Everything else (B2M 3.5-5.5, Albumin any); Stage 3: B2M>5.5^13^.

### Myeloma therapies

A list of currently approved and used anti-myeloma therapies was derived from previously published clinical literature. Treatment with standard anti-myeloma chemotherapeutic regimens for each myeloma patient was assessed using a string search of each cancer therapy in the concept name and manually reviewed for correctness. Bone marrow transplantation/BMT (Hematopoietic Stem Cell Transplantation) was identified using SNOMED code 5960049, which included the vocabulary descendants of the SNOMED codes 42537745 (Bone Marrow Transplant present) and 23719005 (Transplantation of Bone Marrow).

### Severity and Outcome measures

For the purpose of this myeloma patient cohort study, the outcomes of interest were: all-cause mortality (including discharge to hospice) during the index encounter, as well as clinical indicators of severity requiring hospitalization (inpatient/emergency room/intensive care unit/ICU or intensive coronary care unit/ICCU visit), or use of mechanical ventilation (N3C Procedure Concept Set ID 179437741) or extracorporeal membrane oxygenation (ECMO; N3C Procedure Concept Set ID 415149730).

### Statistical analysis and data visualization

All the analyses were performed on the Palantir platform on the N3C data enclave. Summary statistics of descriptive analyses have been represented as counts and percentages of categorical variables. The risk of severe and mild outcomes was calculated using multivariate logistic regression analysis. The models were controlled for age group, gender, race and ethnicity, smoking status, vaccination status, treatment, BMT, and CCI variables. Adjusted odds ratios were estimated with 95% Confidence intervals for potential risk factors. All tests were two-sided. Finally, Cox proportional hazard models with time to death from COVID-19 infection were used to calculate the risk of death, adjusted for age group, gender, race and ethnicity, smoking status, vaccination status, treatment, BMT, and CCI for variables. As per N3C policy, counts of <20 were not reported to privacy.

### Causal effect analysis

We performed matched sample analysis to compute the sample average treatment effect on the treated (SATT) as the measure of the causal effect of the top associated risk factors. The design and development of the SATT method are non-trivial and mathematically involved. For details, please refer to Athey and Imbens 2016^14^. Briefly, let us consider patients *i* who received treatment *T* (for example BMT or vaccination). Let *y*_*i*_ denote the response (for example, <u>probability of death from COVID-19, referred to as “mortality or discharge to hospice” adhering to the spirit of using sensitive language around Covid-related mortality</u>) of patient *i*. The causal effect of the treatment is defined as the difference in the response measure under the condition that the patient received the treatment from the response measure had the patient not received the treatment. Therefore, the causal effect of the treatment on the treated α_*i*_ is defined as

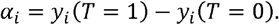

However, in observational data that is not experimentally generated, it is often not possible to observe both the response measures under treatment and no-treatment conditions. For example, for a patient in the dataset that received BMT, we only observe the response under treatment *y*_*i*_(*T*= 1), but we do not observe the response under no-treatment *y*_*i*_(*T*= 0). Let ***x***_i_ denote covariates (such as patient characteristics, disease conditions, etc.) that determine the patients’ likelihood of receiving the treatment. In experimental data, treatments are usually randomized across observation units. However, in observational data, treatments are not usually randomized; rather, treatments are decided based on the covariates that determine both the treatment assignment and the response outcomes. Under the assumption that the treatment assignment is independent of the outcomes given the covariates^14^, that is

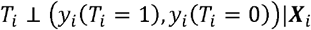

It can be assumed that the response outcome of the patients in the control group can reasonably approximate the response outcome of the patients in the treatment group, given that the patients are matched on the covariates. Therefore, the treatment effect on the treated can be estimated as

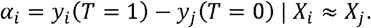

The sample average treatment effect on the treated is then estimated as

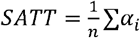

Where *n* is the number of patients who received treatment in the empirical estimation sample, we used a propensity score-based matching. First, we estimated a logistic regression model with the treatment status as the response and the covariates such as age, sex, disease stage and all other relevant variables as explanatory to predict the likelihood of patients receiving the treatment. Then we matched the treatment group with the control group patients by choosing the closest predicted likelihood of receiving the treatment. The SATT is then estimated as the sample average of the difference in the response of the treatment and the control groups.

### The Role of the Institutional Review Board

Prior institutional review board approvals were obtained from respective institutions to access the N3C data. Further, all the authors who had access to N3C data in the Enclave and performed analyses were approved by the N3C data Use Request committee to access the limited use dataset (Level 3).

## RESULTS

As of N3C data release v76 (date 05/06/2022), the N3C database consisted of 1,061,748 cancer patients, out of which 26,064 were myeloma patients (Resource Download Request ID: DRR-DCAB2E1). Among these, 8,588 myeloma patients were COVID-19 positive (**Figure 1: Consort diagram**). In addition, 225 patients had smoldering multiple myeloma (condition: 4184985), while 45 were Monoclonal gammopathy of undetermined significance (MGUS) (conditions: 40297097, 45566693, Observations: 4149022, 37312312, 42511601). We excluded these two subgroups from our analysis. **Table 1** provides detailed patient pre-admission characteristics of our study cohort, including demographic, clinical features, ISS staging, as well as COVID-19 vaccination history. According to N3C policy, cell sizes less than 20 were suppressed using a small cell size indicator (<20) to protect person privacy. To avoid cell sizes being computed from the marginal totals in cases where there is only one small cell in a row or column, we deleted the marginal total for the row or column having four or less elements and retained the small cell size indicator (<20). The mean age at diagnosis of COVID-19-positive patients was 65.89 years. 46.7% were females, and 20.2% were Blacks. 5.79% and 4.88% of myeloma patients were of ISS stages II and III, respectively. 25.56% had a Charlson Comorbidity Index (CCI) score of 0, 12.94% had a score of 1 and 27.54% had a score of 2 or more.

**Table 1:** Preadmission characteristics including demographic, clinical features, co-morbidities, ISS staging, and COVID-19 vaccination status.

|  | **Multiple Myeloma** | **Non-COVID**<br>**17476** | **COVID**<br>**8588** | **Total**<br>**26064** |
| --- | --- | --- | --- | --- |
| **COVID Variant** |  |  |  |  |
|  | Alpha |  | 2553 |  |
|  | Delta |  | 2085 |  |
|  | Omicron |  | 1487 |  |
|  | Unknown |  | 2463 |  |
| **Median Age** |  | 67.65 | 65.89 |  |
| **Gender** |  |  |  |  |
|  | FEMALE | 7942 | 4017 | 11959 |
|  | MALE | 9533 | 4570 | 14103 |
| **Age Group** |  |  |  |  |
|  | 0-17 | <20 | <20 |  |
|  | 18-29 | 66 | 39 | 105 |
|  | 30-49 | 1096 | 700 | 1796 |
|  | 50-64 | 5253 | 2897 | 8150 |
|  | 65+ | 11040 | 4938 | 15978 |
| **Race** |  |  |  |  |
|  | Asian | 331 | 177 | 508 |
|  | Black | 4030 | 1739 | 5769 |
|  | Other | 1742 | 1074 | 2816 |
|  | White | 11290 | 5538 | 16828 |
| **ISS Staging** |  |  |  |  |
|  | Unknown | 11580 | 5252 | 16832 |
|  | Stage I | 5900 | 3023 | 8923 |
|  | Stage II | 953 | 497 | 1450 |
|  | Stage III | 1004 | 419 | 1423 |
| **Smoking Status** |  |  |  |  |
|  | Current or Former | 4560 | 2144 | 6704 |
|  | Non-Smoker | 12878 | 6425 | 19303 |
| **BMI** |  |  |  |  |
|  | Normal weight | 3329 | 1611 | 4940 |
|  | Obese | 4699 | 2243 | 6942 |
|  | Overweight | 4228 | 2081 | 6309 |
|  | Underweight | 307 | 139 | 446 |
| **Renal disease** |  |  |  |  |
|  | No | 10826 | 6269 | 17095 |
|  | Yes | 6612 | 2295 | 8907 |
| **Pulmonary disease** |  |  |  |  |
|  | No | 12521 | 6867 | 19388 |
|  | Yes | 4917 | 1697 | 6614 |
| **CCI Score level** |  |  |  |  |
|  | Mild | 4449 | 2194 | 6643 |
|  | Moderate | 2907 | 1111 | 4018 |
|  | Severe | 6979 | 2365 | 9344 |
| **Diabetes** |  |  |  |  |
|  | No | 11233 | 6235 | 17468 |
|  | Yes | 6185 | 2320 | 8505 |
| **PVD (Peripheral Vascular Disease)** |  |  |  |  |
|  | No | 13834 | 7528 | 21362 |
|  | Yes | 3584 | 1027 | 4611 |
|  |  | **Non-COVID** | **COVID** |  |
| **COVID-19 vaccination** |  |  |  |  |
|  | No | 12829 | 5760 | 18589 |
|  | Yes | 4647 | 2828 | 7475 |
| **Vaccination Type** |  |  |  |  |
|  | JANSSEN | 130 | 68 | 198 |
|  | MODERNA | 1574 | 734 | 2308 |
|  | PFIZER_BIONTECH | 2922 | 1334 | 4256 |
|  | OTHER | 21 | <20 |  |
|  | NONE | 12829 | 5760 | 18589 |

**Figure 1.**
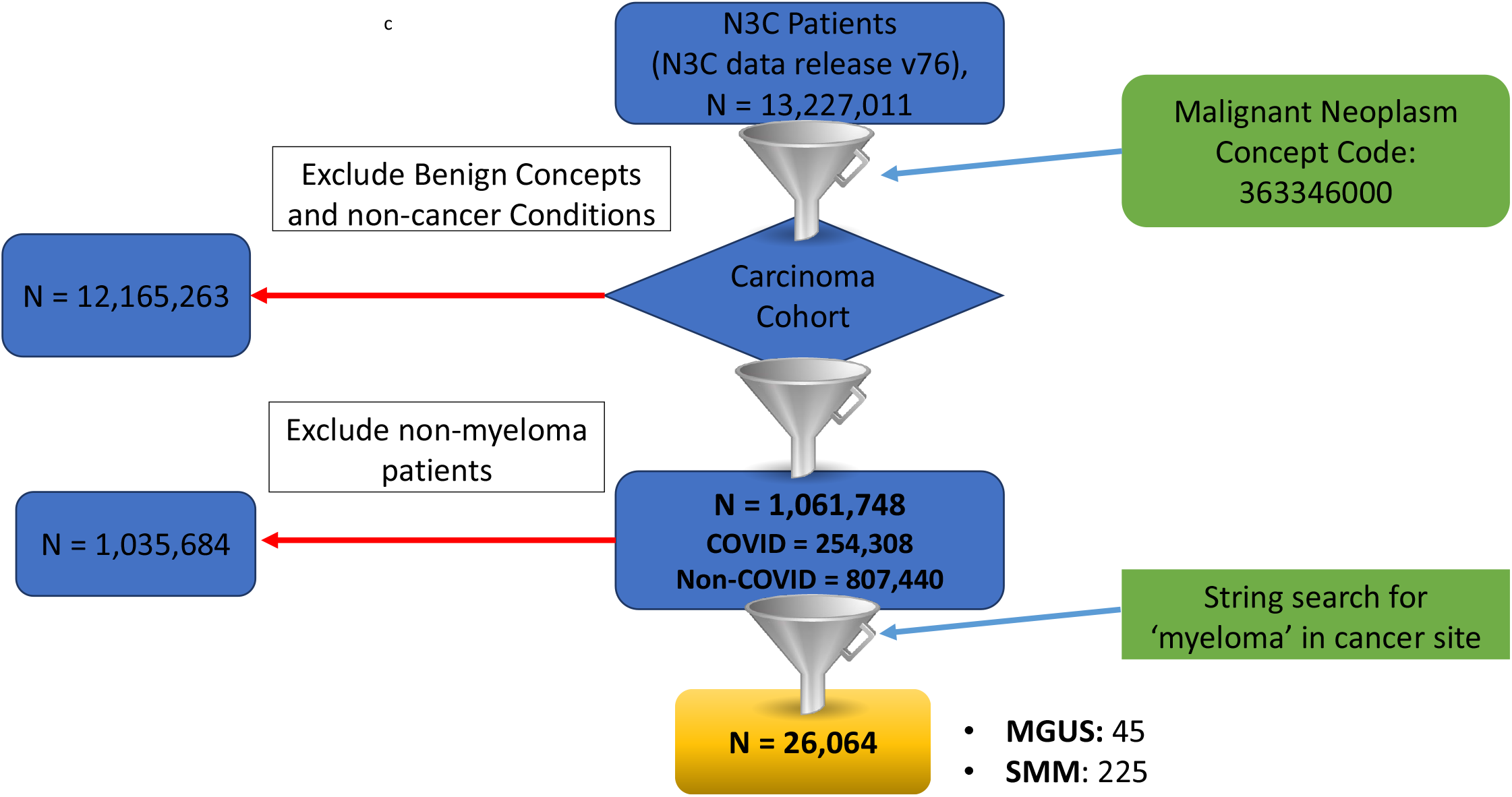
Consort diagram. • Step 1: Use row-level patient data in the N3C Data Enclave, to construct a cohort of patients with myeloma. • Step 2: Analysis of risk factors associated with COVID-19 severity and survival.

Among those with a diagnosis of COVID-19, 3.20% required invasive ventilation, and 55.73% required an inpatient or ED visit. 12.19% of patients underwent Acute kidney injury (AKI) during hospitalization (**Table 2A**). Overall, the survival probability was 90.7% across the course of the study. A total of 1.97% of N3C-myeloma COVID-19-positive patients died within the first 10 days, while 4.47% died in their initial 30 days of COVID-19 hospitalization (**Table 2B**). **Table 3** provides a summary of anti-myeloma medications, including prior bone marrow transplantation. Of the patients with available data, 26.595% had a prior history of blood or marrow transplant (BMT).

**Table 2A:** Summary of severity indicator variables with severity types.

|  |  | Non-COVID | COVID | Total |
| --- | --- | --- | --- | --- |
| **AKI in Hospital** | No | 14701 | 7536 | 22237 |
|  | Yes | 2775 | 1047 | 3822 |
| **Invasive Ventilation** | No | 16852 | 8308 | 25160 |
|  | Yes | 624 | 275 | 899 |
| **Inpatient or ED** | FALSE | 5503 | 3778 | 9281 |
|  | TRUE | 11935 | 4786 | 16721 |

**Table 2B:** Summary of survival indicator variables with survival days.

|  |  | Survival days | % |
| --- | --- | --- | --- |
| **COVID-19 Severity_Type** |  |  |  |
|  | Unaffected | 4522 | 52.65% |
|  | Mild | 1530 | 17.82% |
|  | Mild_ED | 315 | 3.67% |
|  | Moderate | 1570 | 18.28% |
|  | Severe | 77 | 0.90% |
|  | Mortality or discharge to hospice | 574 | 6.68% |
| **Survival_days (COVID-19+ patients)** |  |  |  |
|  | 0-10 | 166 | 1.93% |
|  | 11-20 | 137 | 1.60% |
|  | 21-30 | 81 | 0.94% |
|  | 31+ | 362 | 4.22% |
|  | Died (Non COVID+) | 53 | 0.62% |
|  | Survived | 7790 | 90.71% |
|  | 10-Day survival | 166 | 98.07% |
|  | 30-Day survival | 384 | 95.53% |

**Table 3.** Medication history of COVID-19-positive myeloma patients. Exposure within -/+ 60 days from COVID index date

|  |  | # |
| --- | --- | --- |
| **Lenalidomide/Revlimid** | No | 7154 |
|  | Yes | 1410 |
| **Bortezomib** | No | 8020 |
|  | Yes | 544 |
| **Pomalidomide** | No | 8079 |
|  | Yes | 485 |
| **Carfilzomib** | No | 8362 |
|  | Yes | 202 |
| **Ixazomib** | No | 8374 |
|  | Yes | 190 |
| **Daratumumab** | No | 7902 |
|  | Yes | 662 |
| **Dexamethasone** | No | 3358 |
|  | Yes | 144 |
| **BMT (Bone marrow transplant)** | No | 6304 |
|  | Yes | 2284 |

Results of univariate analyses are shown in **Table 4A**. Categories from Table 1 with <20 patients were subgrouped for logistic regression analysis to obscure/suppress small cell sizes to protect person privacy. At a p-value threshold <0.05 and Odd ratio >1.5, prior history of hypertension (OR=1.90; 95%CI: 1.62-2.23), PVD (OR=1.78; 95%CI: 1.45-2.219), renal disease (OR=2.39; 95%CI: 2.05-2.279), pulmonary disease (OR=2.27; 95%CI: 1.92-2.68) and diabetes (OR=2.08; 95%CI: 1.78-2.44) were significantly correlated with higher COVID-19 severity. Further, CCI score of 2 or more (OR=2.79: 2.30-3.39) and BMI code of ‘underweight’ (<18.5) (OR=2.22: 1.31-3.78) were also associated with higher levels of severity. Race (black vs. white-p<e-99) was highly correlated with severity. On the other hand, vaccination status (OR 0.36: 0.29-0.44) and history of BMT (OR=0.45: 0.37-0.54) showed a protective association with COVID-19 severity. Multivariate logistic regression analysis confirmed the following were found to be associated with higher rates of severity (**Table 4B**): history of pulmonary disease (OR 1.53) and renal disease (OR 1.54), associated with higher risks of severe outcomes. Further, a severe Charlson Comorbidity Index (CCI) score level (OR 1.78) was also associated with an increased risk of COVID-19 severity. Multivariate logistic regression analysis confirmed a negative/protective association between COVID-19 severity with BMT (Adjusted Odds Ratio or AOR 0.79) and COVID-19 vaccination (AOR 0.28). Treatment history of Dexamethasone (AOR 2.23), PI (AOR 1.5), and IMiD (AOR 1.4) therapy was found significantly correlated with an increase in the risk of COVID-19 severity **Table 6** provides a summary of multivariate cox proportional regression analysis in the myeloma-COVID-19-positive cohort. CCI score high (Cox Hazards’ ratio/ HR 5.2; 95%CI: 2.55-10.36), CCI score moderate (HR 3.34; 95%CI: 1.68-6.65) ISS moderate or severe stage (HR 1.55; 95%CI: 0.98-2.5), history of PVD (OR 1.62; 95% 1.12-2.34), and diabetes were highly significantly correlated with poorer patient survival. In addition, proteasome inhibitor treatment (HR 1.6; 95%CI: 1.1-2.5) was also significantly associated. On the other hand, BMT (HR 0.65; 95%CI: 0.42-1) and COVID-19 vaccination (HR 0.302; 95%CI: 0.19-0.47) were associated with a significantly lower risk of death and higher survival following COVID-19 infection in myeloma patients.

**Table 4A:**
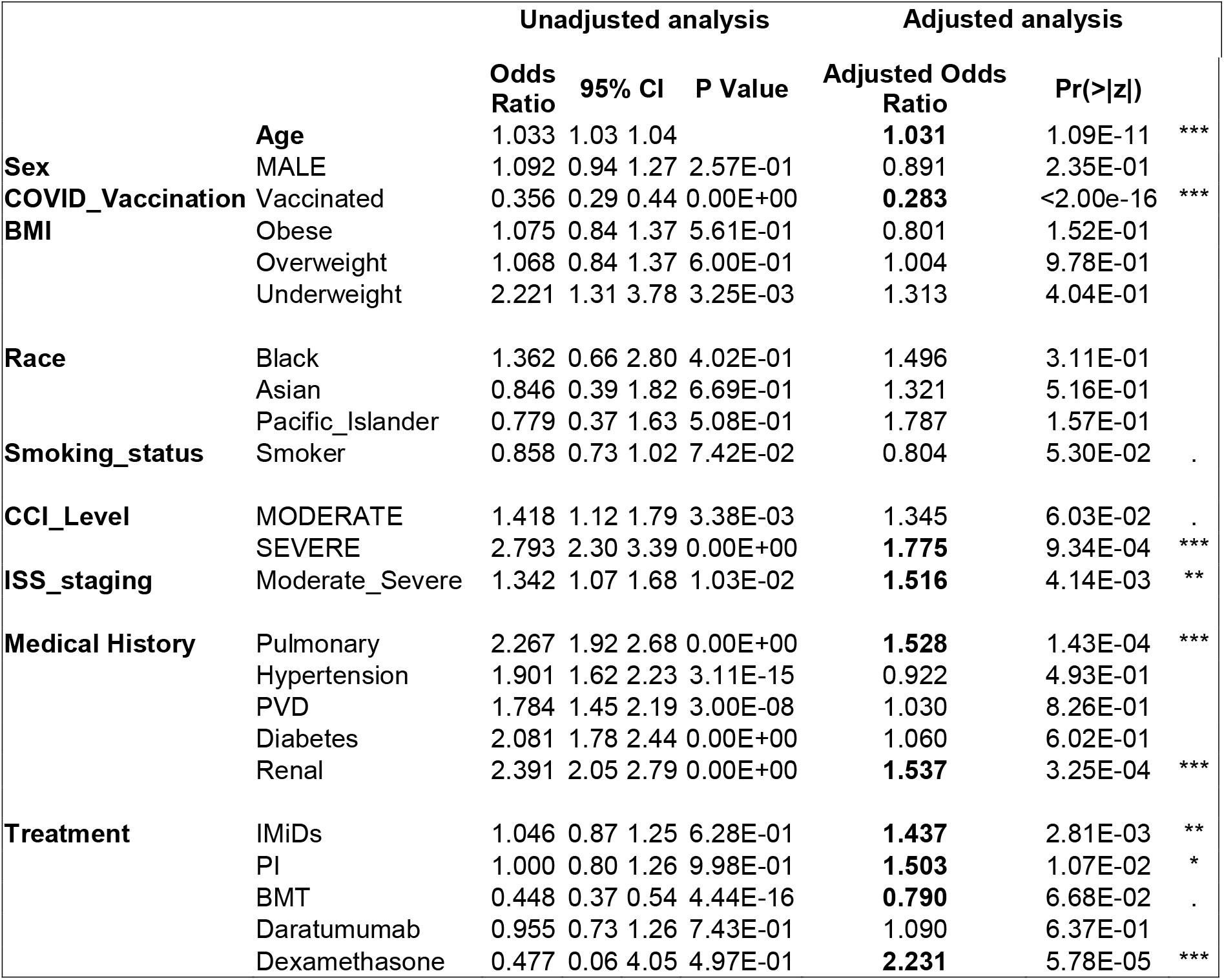
Multivariate logistic regression analysis results (association with severity)

**Table 4B:** Multivariate Cox regression analysis results (association with survival)

|  |  | Adjusted Hazards<br>Ratio (coef) | lower 0.95 | upper 0.95 | Pr(>\|z\|) |  |
| --- | --- | --- | --- | --- | --- | --- |
| **Age** |  | **1.040** | 1.02 | 1.06 | 3.98E-07 | *** |
| **Sex** | MALE | 0.793 | 0.57 | 1.09 | 1.58E-01 |  |
| **COVID Vaccination** | Vaccinated | **0.302** | 0.19 | 0.47 | 1.02E-07 | *** |
| **BMI** | Obese | 1.045 | 0.67 | 1.64 | 8.48E-01 |  |
|  | Overweight | 1.287 | 0.84 | 1.97 | 2.44E-01 |  |
|  | Underweight | 0.808 | 0.28 | 2.31 | 6.90E-01 |  |
| **Race** | Black | 2.630 | 0.36 | 19.21 | 3.41E-01 |  |
|  | Asian | 2.192 | 0.27 | 17.72 | 4.62E-01 |  |
|  | Pacific_Islander | 2.847 | 0.38 | 21.24 | 3.08E-01 |  |
| **Smoking status** | Smoker | 0.918 | 0.65 | 1.30 | 6.27E-01 |  |
| **CCI Level** | MODERATE | **3.340** | 1.68 | 6.65 | 6.05E-04 | *** |
|  | SEVERE | **5.138** | 2.55 | 10.36 | 4.84E-06 | *** |
| **ISS staging** | Moderate_Severe | **1.549** | 0.98 | 2.45 | 6.22E-02 | . |
| **Treatment** | Pulmonary | 1.009 | 0.71 | 1.43 | 9.62E-01 |  |
|  | Hypertension | 0.961 | 0.64 | 1.45 | 8.49E-01 |  |
|  | PVD | **1.619** | 1.12 | 2.34 | 1.05E-02 | * |
|  | Diabetes | **0.737** | 0.52 | 1.04 | 8.29E-02 | . |
|  | Renal | 1.080 | 0.75 | 1.55 | 6.80E-01 |  |
| **Medical History** | IMiDs | 1.257 | 0.87 | 1.82 | 2.28E-01 |  |
|  | PI | **1.623** | 1.06 | 2.48 | 2.46E-02 | * |
|  | BMT | **0.653** | 0.42 | 1 | 5.49E-02 | . |
|  | Daratumumab | 1.291 | 0.80 | 2.09 | 2.97E-01 |  |

Finally, we performed causal estimation using matched sample SATT method as detailed in the Methods section. A matched sample analysis follows two steps^14^. In the first step, for every individual patient in the treatment group (for example, patients who received BMT and/or vaccination), a sub-sample of patients from the control group (for example, patients who did not receive either BMT or vaccination) who are similar to the treatment group patient in every aspect other than the treatment (BMT or vaccination). The difference in the survival probability or duration (or any other relevant response measure) between the patient in the treatment group and the matched patient in the control group is the treatment effect. The average difference in the response measure between the patients in the treatment group and the control group is the SATT. Our causal effect analysis confirmed that BMT and vaccination status were associated with decreased risk of COVID-19-related death in myeloma patients, while the history of pulmonary disease, renal disease, as well as IMiD and PI therapy was significantly correlated with a high risk of death. The SATT of BMT as treatment and probability of survival status (death = 1) as the response is -0.025 (−0.031, -0.020). This indicates that MM patients who received BMT treatment are significantly less likely to die from COVID-19 as compared to those MM patients who did not receive BMT. Similarly, and not surprisingly SATT for vaccination status is -0.123 (−0.127, -0.118) with a Welch t-Statistic of -55.186 (p-Value < 2.2e-16). This indicates that MM patients who received vaccination are significantly less likely to die from COVID-19 than patients who did not receive the vaccination. Interestingly, the pre-existence of pulmonary and renal complications significantly increases the chances of death from COVID-19.

## DISCUSSION

We have used the N3C patient cohort that currently includes > 8 million COVID-tested patients with at least 1 clinical encounter at >75 US medical centers to construct a cohort of COVID-19 patients with multiple myeloma. To the best of our knowledge, this is the largest nationwide study on multiple myeloma patients with COVID-19 infection.

We identified several known and so-far unknown characteristics as potential risk factors for severity and death in multiple myeloma. For example, several groups, including us, have earlier established the impact of male gender and existing comorbidities as risk factors associated with COVID-19.

The impact of race on COVID-19-associated mortality/severity has been controversial. Although some studies have shown racial disparities to be significantly associated with mortality risk, several others did not find any significant effect on the rate of hospitalization or mortality. We observed significantly higher risk associated with severity in non-white ethnic groups compared to whites. These results require further in-depth analysis exploring social determinants of health, socioeconomic parameters, and access to timely and appropriate healthcare.

Furthermore, interestingly, age was not found to be significantly associated with either severity or death in our N3C-myeloma cohort. This was probably since the median age of diagnosis was already above 65 years, which has been shown as the at-risk age earlier.

We showed that vaccination with two doses of Pfizer or Moderna vaccine or a single-dose of J&J vaccine showed a protective effect in the N3C-myeloma cohort. Vaccinated myeloma patients were at >350% less risk of severe outcomes and 331% less risk of death following COVID-19 infection compared to unvaccinated myeloma patients. An earlier study demonstrated that 2/3^rd^ of vaccinated myeloma patients show some response to mRNA vaccines, although vaccination may only provide partial protection from infection, while 1/3^rd^ failed to respond based on background IgG levels of 50IU/ml^15^. However, the threshold/cut-offs were primarily probabilistic, with no clinical follow-up correlating relevant anti-spike IgG levels with protection in vaccinated patients. A recent study that measured vaccine-induced neutralizing antibodies (nAbs) in myeloma patients receiving SARS CoV-2 vaccination found that, although more than 80% of patients showed serological response to vaccines, several patients lack detectable virus-neutralizing activity for protection from COVID-19 which was affected by race, disease state, treatment, etc. Therefore, for a reliable evaluation of immunogenicity of COVID-19 vaccines in myeloma patients, regular management and monitoring of nAbs titer and SARS CoV-2 is crucial^16,17^.

Next, we found that bone marrow transplant (BMT) has more than 1.5 folds protective effect on both severity and death. On the other hand, an earlier study showed that patients with COVID-19 (including 90 patients with multiple myeloma with a prior history of autologous and allogeneic hematopoietic stem-cell transplantation (HSCT) have poor overall survival^18^. Interestingly, a recent study by Romano *et al*. (2022) showed that absolute monocyte count prior to SARS-CoV-2 infection is predictive of the risk of overall survival in patients with heme malignancies^19^.

Next, we focused on drug classes commonly used as anti-myeloma therapies. Proteasome inhibitors (PIs) are standard-of-care/primary chemotherapeutic agents for myeloma^20–24^. Bortezomib (Bz/Velcade) was the first proteasome inhibitor to be approved by the US Food and Drug Administration (FDA) for clinical application in 2003 for the treatment of relapsed and refractory myeloma^5,25,26^. Other FDA-approved second-generation proteasome inhibitors used as anti-myeloma drugs include carfilzomib (Cz/Kyprolis) and the oral medication ixazomib (Ix /Ninlaro/MLN9708)^25–27^. PIs are effective anti-MM drugs when used alone or in combination with other anti-cancer agents like immunomodulatory drugs (IMiDs), alkylating agents, topoisomerase inhibitors, corticosteroids, and histone deacetylase inhibitors (HDACis)^5,21^. More recent improvements in anti-myeloma therapeutic regimens include the monoclonal antibody Daratumumab (targeting CD-38) and chimeric antigen receptor or CAR-T-cell therapy.

Very interestingly, both our univariate and multivariate analysis showed that treatment with Immunomodulatory agents (Lenalidomide, Revlimid and Pomalidomide) were significantly associated with severe outcomes and all-cause mortality. The risk of severe outcomes was two-fold IMiDs, while the risk of death was more than 2.5 folds in myeloma patients on IMiDs compared to the patients who were not on IMiDs during the study period.

Finally, anti-myeloma monoclonal antibody therapy was found highly protective in COVID-19-affected myeloma patients. The risk of severity was 50% lower in patients treated with daratumumab compared to patients being administered other ant-myeloma therapies. The correlation between daratumumab and IMiD-based systemic therapy, resultant immunoparesis or compromised immune system and severe/adverse COVID-19 outcomes have so far been conflicting^28,29^. However, most of these studies were not powered enough. Therefore, we suggest careful clinical monitoring and treatment for the management of myeloma patients with COVID-19 for immune system dysregulation during disease progression and/or immunomodulatory therapies.

Since causal effect analysis models demonstrate the ‘cause’ from a statistical standpoint, the determination of the actual biological causes needs further clinical research that compares each of these risk factors. Furthermore, a biomarker analysis, including characterization of immune and inflammatory cell populations as well as pro-inflammatory cytokines in patients with MM, will be necessary.

Nevertheless, our analysis method may serve as a template for identifying major risk factors associated with death and severity in future pandemic scenarios using large-scale patient-centered databases.

We have earlier elaborated on the strengths of the N3C database, its comparability with the manually extracted registry data from the CCC19 cohort, as well as our mechanisms to perform strict data QC, as well as the weaknesses related to the heterogeneity in data collection & reporting processes at various hospital systems, data portability, and data missingness^12^. With progressive changes within the N3C cohort and the development of more and better quality tools to extract and harmonize data, we have been able to create a robust dataset of COVID-19 patients and non-COVID-19 controls in our database.

Overall, through the creation of the N3C-myeloma dataset, the largest COVID-19 and multiple myeloma cohort in the United States reported so far, this article summarizes the risk of severe outcomes and death/all-cause mortality associated with COVID-19 patients in multiple myeloma. Our cohort provides us several options to perform large-scale observational studies comparing various vaccination schedules, as well as differences between severity and survival between COVID-19 variants of concern, like delta vs. Omicron.

## Data Availability

All data produced in the present work are contained in the manuscript.

## AUTHOR CONTRIBUTIONS

“Authorship was determined using ICMJE recommendations.”

## ^¥^Consortial contributors

Christopher G Chute^1^, Richard A Moffitt^2^, Melissa A Haendel^3^

1. Johns Hopkins University, Baltimore, MD, USA.
2. Department of Biomedical Informatics, Stony Brook University, Stony Brook, NY.
3. Center for Health AI, University of Colorado School of Medicine, Aurora, CO, USA.

## N3C ATTRIBUTION

The analyses described in this publication were conducted with data or tools accessed through the NCATS N3C Data Enclave covid.cd2h.org/enclave and supported by CD2H - The National COVID Cohort Collaborative (N3C) IDeA CTR Collaboration 3U24TR002306-04S2 NCATS U24 TR002306. This research was possible because of the patients whose information is included within the data from participating organizations (covid.cd2h.org/dtas) and the organizations and scientists (covid.cd2h.org/duas) who have contributed to the ongoing development of this community resource (cite this https://doi.org/10.1093/jamia/ocaa196).

## Disclaimer

The content is solely the responsibility of the authors and does not necessarily represent the official views of the National Institutes of Health or the N3C program.

## IRB

The N3C data transfer to NCATS is performed under a Johns Hopkins University Reliance Protocol # IRB00249128 or individual site agreements with NIH. The N3C Data Enclave is managed under the authority of the NIH; information can be found at https://ncats.nih.gov/n3c/resources.

## INDIVIDUAL ACKNOWLEDGEMENTS FOR CORE CONTRIBUTORS

We gratefully acknowledge the Oncology Domain Team and the following core contributors to N3C: Adam B. Wilcox, Adam M. Lee, Alexis Graves, Alfred (Jerrod) Anzalone, Amin Manna, Amit Saha, Amy Olex, Andrea Zhou, Andrew E. Williams, Andrew Southerland, Andrew T. Girvin, Anita Walden, Anjali A. Sharathkumar, Benjamin Amor, Benjamin Bates, Brian Hendricks, Brijesh Patel, Caleb Alexander, Carolyn Bramante, Cavin Ward-Caviness, Charisse Madlock-Brown, Christine Suver, Christopher Chute, Christopher Dillon, Chunlei Wu, Clare Schmitt, Cliff Takemoto, Dan Housman, Davera Gabriel, David A. Eichmann, Diego Mazzotti, Don Brown, Eilis Boudreau, Elaine Hill, Elizabeth Zampino, Emily Carlson Marti, Emily R. Pfaff, Evan French, Farrukh M Koraishy, Federico Mariona, Fred Prior, George Sokos, Greg Martin, Harold Lehmann, Heidi Spratt, Hemalkumar Mehta, Hongfang Liu, Hythem Sidky, J.W. Awori Hayanga, Jami Pincavitch, Jaylyn Clark, Jeremy Richard Harper, Jessica Islam, Jin Ge, Joel Gagnier, Joel H. Saltz, Joel Saltz, Johanna Loomba, John Buse, Jomol Mathew, Joni L. Rutter, Julie A. McMurry, Justin Guinney, Justin Starren, Karen Crowley, Katie Rebecca Bradwell, Kellie M. Walters, Ken Wilkins, Kenneth R. Gersing, Kenrick Dwain Cato, Kimberly Murray, Kristin Kostka, Lavance Northington, Lee Allan Pyles, Leonie Misquitta, Lesley Cottrell, Lili Portilla, Mariam Deacy, Mark M. Bissell, Marshall Clark, Mary Emmett, Mary Morrison Saltz, Matvey B. Palchuk, Melissa A. Haendel, Meredith Adams, Meredith Temple-O’Connor, Michael G. Kurilla, Michele Morris, Nabeel Qureshi, Nasia Safdar, Nicole Garbarini, Noha Sharafeldin, Ofer Sadan, Patricia A. Francis, Penny Wung Burgoon, Peter Robinson, Philip R.O. Payne, Rafael Fuentes, Randeep Jawa, Rebecca Erwin-Cohen, Rena Patel, Richard A. Moffitt, Richard L. Zhu, Rishi Kamaleswaran, Robert Hurley, Robert T. Miller, Saiju Pyarajan, Sam G. Michael, Samuel Bozzette, Sandeep Mallipattu, Satyanarayana Vedula, Scott Chapman, Shawn T. O’Neil, Soko Setoguchi, Stephanie S. Hong, Steve Johnson, Tellen D. Bennett, Tiffany Callahan, Umit Topaloglu, Usman Sheikh, Valery Gordon, Vignesh Subbian, Warren A. Kibbe, Wenndy Hernandez, Will Beasley, Will Cooper, William Hillegass, Xiaohan Tanner Zhang. Details of contributions available at covid.cd2h.org/core-contributors

## DATA PARTNERS WITH RELEASED DATA

The following institutions whose data is released or pending:

Available: Advocate Health Care Network — UL1TR002389: The Institute for Translational Medicine (ITM) • Boston University Medical Campus — UL1TR001430: Boston University Clinical and Translational Science Institute • Brown University — U54GM115677: Advance Clinical Translational Research (Advance-CTR) • Carilion Clinic — UL1TR003015: iTHRIV Integrated Translational health Research Institute of Virginia • Charleston Area Medical Center — U54GM104942: West Virginia Clinical and Translational Science Institute (WVCTSI) • Children’s Hospital Colorado — UL1TR002535: Colorado Clinical and Translational Sciences Institute • Columbia University Irving Medical Center — UL1TR001873: Irving Institute for Clinical and Translational Research • Duke University — UL1TR002553: Duke Clinical and Translational Science Institute • George Washington Children’s Research Institute — UL1TR001876: Clinical and Translational Science Institute at Children’s National (CTSA-CN) • George Washington University — UL1TR001876: Clinical and Translational Science Institute at Children’s National (CTSA-CN) • Indiana University School of Medicine — UL1TR002529: Indiana Clinical and Translational Science Institute • Johns Hopkins University — UL1TR003098: Johns Hopkins Institute for Clinical and Translational Research • Loyola Medicine — Loyola University Medical Center • Loyola University Medical Center — UL1TR002389: The Institute for Translational Medicine (ITM) • Maine Medical Center — U54GM115516: Northern New England Clinical & Translational Research (NNE-CTR) Network • Massachusetts General Brigham — UL1TR002541: Harvard Catalyst • Mayo Clinic Rochester — UL1TR002377: Mayo Clinic Center for Clinical and Translational Science (CCaTS) • Medical University of South Carolina — UL1TR001450: South Carolina Clinical & Translational Research Institute (SCTR) • Montefiore Medical Center — UL1TR002556: Institute for Clinical and Translational Research at Einstein and Montefiore • Nemours — U54GM104941: Delaware CTR ACCEL Program • NorthShore University HealthSystem — UL1TR002389: The Institute for Translational Medicine (ITM) • Northwestern University at Chicago — UL1TR001422: Northwestern University Clinical and Translational Science Institute (NUCATS) • OCHIN — INV-018455: Bill and Melinda Gates Foundation grant to Sage Bionetworks • Oregon Health & Science University — UL1TR002369: Oregon Clinical and Translational Research Institute • Penn State Health Milton S. Hershey Medical Center — UL1TR002014: Penn State Clinical and Translational Science Institute • Rush University Medical Center — UL1TR002389: The Institute for Translational Medicine (ITM) • Rutgers, The State University of New Jersey — UL1TR003017: New Jersey Alliance for Clinical and Translational Science • Stony Brook University — U24TR002306 • The Ohio State University — UL1TR002733: Center for Clinical and Translational Science • The State University of New York at Buffalo — UL1TR001412: Clinical and Translational Science Institute • The University of Chicago — UL1TR002389: The Institute for Translational Medicine (ITM) • The University of Iowa — UL1TR002537: Institute for Clinical and Translational Science • The University of Miami Leonard M. Miller School of Medicine — UL1TR002736: University of Miami Clinical and Translational Science Institute • The University of Michigan at Ann Arbor — UL1TR002240: Michigan Institute for Clinical and Health Research • The University of Texas Health Science Center at Houston — UL1TR003167: Center for Clinical and Translational Sciences (CCTS) • The University of Texas Medical Branch at Galveston — UL1TR001439: The Institute for Translational Sciences • The University of Utah — UL1TR002538: Uhealth Center for Clinical and Translational Science • Tufts Medical Center — UL1TR002544: Tufts Clinical and Translational Science Institute • Tulane University — UL1TR003096: Center for Clinical and Translational Science • University Medical Center New Orleans — U54GM104940: Louisiana Clinical and Translational Science (LA CaTS) Center • University of Alabama at Birmingham — UL1TR003096: Center for Clinical and Translational Science • University of Arkansas for Medical Sciences — UL1TR003107: UAMS Translational Research Institute • University of Cincinnati — UL1TR001425: Center for Clinical and Translational Science and Training • University of Colorado Denver, Anschutz Medical Campus — UL1TR002535: Colorado Clinical and Translational Sciences Institute • University of Illinois at Chicago — UL1TR002003: UIC Center for Clinical and Translational Science • University of Kansas Medical Center — UL1TR002366: Frontiers: University of Kansas Clinical and Translational Science Institute • University of Kentucky — UL1TR001998: UK Center for Clinical and Translational Science • University of Massachusetts Medical School Worcester — UL1TR001453: The UMass Center for Clinical and Translational Science (UMCCTS) • University of Minnesota — UL1TR002494: Clinical and Translational Science Institute • University of Mississippi Medical Center — U54GM115428: Mississippi Center for Clinical and Translational Research (CCTR) • University of Nebraska Medical Center — U54GM115458: Great Plains IDeA-Clinical & Translational Research • University of North Carolina at Chapel Hill — UL1TR002489: North Carolina Translational and Clinical Science Institute • University of Oklahoma Health Sciences Center — U54GM104938: Oklahoma Clinical and Translational Science Institute (OCTSI) • University of Rochester — UL1TR002001: UR Clinical & Translational Science Institute • University of Southern California — UL1TR001855: The Southern California Clinical and Translational Science Institute (SC CTSI) • University of Vermont — U54GM115516: Northern New England Clinical & Translational Research (NNE-CTR) Network • University of Virginia — UL1TR003015: iTHRIV Integrated Translational health Research Institute of Virginia • University of Washington — UL1TR002319: Institute of Translational Health Sciences • University of Wisconsin-Madison — UL1TR002373: UW Institute for Clinical and Translational Research • Vanderbilt University Medical Center — UL1TR002243: Vanderbilt Institute for Clinical and Translational Research • Virginia Commonwealth University — UL1TR002649: C. Kenneth and Dianne Wright Center for Clinical and Translational Research • Wake Forest University Health Sciences — UL1TR001420: Wake Forest Clinical and Translational Science Institute • Washington University in St. Louis — UL1TR002345: Institute of Clinical and Translational Sciences • Weill Medical College of Cornell University — UL1TR002384: Weill Cornell Medicine Clinical and Translational Science Center • West Virginia University — U54GM104942: West Virginia Clinical and Translational Science Institute (WVCTSI)

Submitted: Icahn School of Medicine at Mount Sinai — UL1TR001433: ConduITS Institute for Translational Sciences • The University of Texas Health Science Center at Tyler — UL1TR003167: Center for Clinical and Translational Sciences (CCTS) • University of California, Davis — UL1TR001860: UCDavis Health Clinical and Translational Science Center • University of California, Irvine — UL1TR001414: The UC Irvine Institute for Clinical and Translational Science (ICTS) • University of California, Los Angeles — UL1TR001881: UCLA Clinical Translational Science Institute • University of California, San Diego — UL1TR001442: Altman Clinical and Translational Research Institute • University of California, San Francisco — UL1TR001872: UCSF Clinical and Translational Science Institute

Pending: Arkansas Children’s Hospital — UL1TR003107: UAMS Translational Research Institute • Baylor College of Medicine — None (Voluntary) • Children’s Hospital of Philadelphia — UL1TR001878: Institute for Translational Medicine and Therapeutics • Cincinnati Children’s Hospital Medical Center — UL1TR001425: Center for Clinical and Translational Science and Training • Emory University — UL1TR002378: Georgia Clinical and Translational Science Alliance • HonorHealth — None (Voluntary) • Loyola University Chicago — UL1TR002389: The Institute for Translational Medicine (ITM) • Medical College of Wisconsin — UL1TR001436: Clinical and Translational Science Institute of Southeast Wisconsin • MedStar Health Research Institute — UL1TR001409: The Georgetown-Howard Universities Center for Clinical and Translational Science (GHUCCTS) • MetroHealth — None (Voluntary) • Montana State University — U54GM115371: American Indian/Alaska Native CTR • NYU Langone Medical Center — UL1TR001445: Langone Health’s Clinical and Translational Science Institute • Ochsner Medical Center — U54GM104940: Louisiana Clinical and Translational Science (LA CaTS) Center • Regenstrief Institute — UL1TR002529: Indiana Clinical and Translational Science Institute • Sanford Research — None (Voluntary) • Stanford University — UL1TR003142: Spectrum: The Stanford Center for Clinical and Translational Research and Education • The Rockefeller University — UL1TR001866: Center for Clinical and Translational Science • The Scripps Research Institute — UL1TR002550: Scripps Research Translational Institute • University of Florida — UL1TR001427: UF Clinical and Translational Science Institute • University of New Mexico Health Sciences Center — UL1TR001449: University of New Mexico Clinical and Translational Science Center • University of Texas Health Science Center at San Antonio — UL1TR002645: Institute for Integration of Medicine and Science • Yale New Haven Hospital — UL1TR001863: Yale Center for Clinical Investigation

