## Supplemental Table 1 for "Sample average treatment effect on the treated analysis using counterfactual explanation identifies BMT and SARS-CoV-2 vaccination as protective risk factors associated with COVID-19 severity and survival in patients with multiple myeloma"

**Supplementary Table 1: Key variables and concept definitions**

| **Variable** |  | | **Concepts and Logic** |
| --- | --- | --- | --- |
|  | | ***Key Outcome*** | |
| ***COVID-19 definition*** |  | | **Concept Set Names (latest codeset ID) with Description:**  N3C Covid diagnosis (35486128)  *Description: Corresponds to ICD10CM Code: U07.1 (condition). Includes 1 Concept (Disease caused by 2019-nCoV - Concept Id: 840539006).*  ATLAS SARS-CoV-2 rt-PCR and AG (651620200)  *Description: Includes 55 Concepts (Measurements), characterizing a positive PCR or Antigen test.*  Atlas #818 [N3C] CovidAntibody retry (45478367)  *Description: Includes 24 Concepts (Measurements), characterizing a positive Antibody test.*  ResultsPos (400691529)  *Description: Includes 6 Concepts , collecting all the affirmative measurement results*  **Logic:**  Patient record must be associated with a “N3C Covid diagnosis”  OR  have at least one of the covid measurement concepts (“ATLAS SARS-CoV-2 rt-PCR and AG” or “[N3C] CovidAntibody retry”) AND a positive result (“ResultsPos”) |
|  | | ***Key Exposures*** | |
| Carcinoma |  | | 45029076 Neoplasm_ODT  **Logic:**  Patients with malignant neoplasm. |
| Multiple Myeloma (MM) |  | | **Logic:**  Any patient with cancer site like '%myeloma%' |
| smoldering multiple myeloma |  | | **Logic:**  Any carcinoma patient with condition concept ID 4184985 |
| Monoclonal gammopathy of undetermined significance |  | | **Logic:**  Any carcinoma patient with condition concept ID 40297097, 45566693  Any carcinoma patient with observation concept ID 4149022, 37312312, 42511601 |
|  | | ***COVID-19 Disease Severity*** | |
| Emergency Department visit |  | | **Visit Concept IDs and Names:**  262 Emergency Room and Inpatient Visit  9203 Emergency Room Visit  **Logic:**  No concept sets were used in this definition.  Includes patients with a visit start date between 14 days prior to the earliest covid diagnosis to 45 days after covid diagnosis that are also associated with one of the listed visit concept IDs. |
| Hospitalization |  | | **Visit Concept IDs and Names:**  262 Emergency Room and Inpatient Visit  8717 Inpatient Hospital  9201 Inpatient Visit  581379 Inpatient Critical Care Facility  **Logic:**  No concept sets were used in this definition. Includes patients with a visit start date between 14 days prior to the earliest covid diagnosis to 45 days after covid diagnosis that are also associated with one of the listed visit concept IDs. |
| Invasive ventilation |  | | **Concept Set Names (latest codeset ID):**  Invasive Mechanical Ventilation 2OCT20 (179437741)  **Logic:**  Includes patients associated with any procedure codes in the concept sets listed that had a procedure date on or after the earliest covid diagnosis. |
| ECMO |  | | **Concept Set Names (latest codeset ID):**  Kostka - ECMO (415149730)  **Logic:**  Includes patients associated with any procedure codes in the concept sets listed that had a procedure date on or after the earliest covid diagnosis. |
| Death |  | | **Logic:**  No concepts or concept sets were used.  Patients were flagged as deceased if a valid entry with a date was included in the death table, which is part of the OMOP data model used in the Enclave. |
|  | | ***Comorbid Conditions*** | |
| Severe cardiovascular event (Congestive heart failure and myocardial infarction) |  | | **Concept Set Names (latest codeset ID):**  Charlson - CHF (359043664)  Charlson - MI (259495957)  **Logic:**  Includes patients associated with any condition codes in the concept sets listed with an occurrence date before the earliest covid diagnosis. |
| Peripheral vascular diseases |  | | **Concept Set Names (latest codeset ID):**  Charlson - PVD (376881697)  **Logic:**  Includes patients associated with any condition codes in the concept sets listed with an occurrence date before the earliest covid diagnosis. |
| Stroke |  | | **Concept Set Names (latest codeset ID):**  Charlson - Stroke (652711186)  **Logic:**  Includes patients associated with any condition codes in the concept sets listed with an occurrence date before the earliest covid diagnosis. |
| Dementia |  | | **Concept Set Names (latest codeset ID):**  Charlson - Dementia (78746470)  **Logic:**  Includes patients associated with any condition codes in the concept sets listed with an occurrence date before the earliest covid diagnosis. |
| Pulmonary Diseases |  | | **Concept Set Names (latest codeset ID):**  Charlson - Pulmonary (514953976)  **Logic:**  Includes patients associated with any condition codes in the concept sets listed with an occurrence date before the earliest covid diagnosis. |
| Rheumatic Diseases |  | | **Concept Set Names (latest codeset ID):**  Charlson - Rheumatic (765004404)  **Logic:**  For all RA patients this value is set to 1. |
| Peptic ulcer diseases |  | | **Concept Set Names (latest codeset ID):**  Charlson - PUD (510748896)  **Logic:**  Includes patients associated with any condition codes in the concept sets listed with an occurrence date before the earliest covid diagnosis. |
| Liver diseases (mild and severe liver diseases) |  | | **Concept Set Names (latest codeset ID):**  Charlson - LiverMild (494981955)  Charlson - LiverSevere (248333963)  **Logic:**  Includes patients associated with any condition codes in the concept sets listed with an occurrence date before the earliest covid diagnosis. |
| Diabetes mellitus (diabetes mellitus and diabetes mellitus with complications) |  | | **Concept Set Names (latest codeset ID):**  Charlson - DM (719585646)  Charlson - DMcx (403438288)  **Logic:**  Includes patients associated with any condition codes in the concept sets listed with an occurrence date before the earliest covid diagnosis. |
| Renal diseases |  | | **Concept Set Names (latest codeset ID):**  Charlson - Renal (220495690)  **Logic:**  Includes patients associated with any condition codes in the concept sets listed with an occurrence date before the earliest covid diagnosis. |
| Cancer (metastatic and non-metastatic) |  | | **Concept Set Names (latest codeset ID):**  Charlson - Cancer (535274723)  Charlson - Mets (378462283)  **Logic:**  Includes patients associated with any condition codes in the concept sets listed with an occurrence date before the earliest covid diagnosis. |
|  |  | | ***COVID-19 Risk factors not included in Deyo-Charlson*** |
| Hypertension |  | | **Concept Set Names (latest codeset ID):**  [LEGEND]Hypertension - 146797511 |
| Coronary artery disease |  | | **Concept Set Names (latest codeset ID):**  Coronary Artery DiseaseV2 Atlas 811 - 630858234 |
|  |  | | ***MM Medications*** |
| tocilizumab |  | | **Logic:**  Includes patients associated with any drug concept IDs where concept name like ‘%tocilizumab%’ |
| ibrutinib |  | | **Logic:**  Includes patients associated with any drug concept IDs where concept name like ‘%ibrutinib%’ |
| lenalidomide |  | | **Logic:**  Includes patients associated with any drug concept IDs where concept name like ‘%lenalidomide%’ |
| revlimid |  | | **Logic:**  Includes patients associated with any drug concept IDs where concept name like ‘%revlimid%’ |
| Pomalidomide |  | | **Logic:**  Includes patients associated with any drug concept IDs where concept name like ‘%pomalidomide%’ |
| Bortezomib |  | | **Logic:**  Includes patients associated with any drug concept IDs where concept name like ‘%bortezomib%’ |
| Carfilzomib |  | | **Logic:**  Includes patients associated with any drug concept IDs where concept name like ‘%carfilzomib%’ |
| Ixazomib |  | | **Logic:**  Includes patients associated with any drug concept IDs where concept name like ‘%ixazomib%’ |
| Daratumumab |  | | **Logic:**  Includes patients associated with any drug concept IDs where concept name like ‘%daratumumab%’ |
| Selinexor |  | | **Logic:**  Includes patients associated with any drug concept IDs where concept name like ‘%selinexor%’ |
| Panobinostat |  | | **Logic:**  Includes patients associated with any drug concept IDs where concept name like ‘%panobinostat%’ |
|  |  | | ***COVID Medications*** |
|  |  | | **Concept Set Names (latest codeset ID):**  Amiodarone gtt 477011432  Anakinra 614076948  Azithromycin 359938251  Chloroquine 818210864  Dexamethasone 213873961  dialysis CRRT/HD 62297511  Dobutamine gtt 41638290  Dopamine gtt 89980583  Epinephrine gtt 138921458  Epoprostenol 820810867  Esmolol gtt 437105398  Hydrocortisone 687120559  Hydroxychloroquine 726349556  Inhaled Nitric Oxide 285273342  intravenous immunoglobulin 241604784  Isoproterenol gtt 514747904  Levosimendan gtt 220815705  Lopinavir 435362039  Lopinavir/Ritonavir combination 165611849  Methylprednisolone 302593795  Milrinone gtt 316286704  Norepinephrine gtt 9512899  Phenylephrine gtt 226615355  Plasma 45129226  Prednisolone 804783116  Prednisone 520650412  Remdesivir 719693192  Ritonavir 407316475  Tocilizumab 889879486  Vasopressin gtt 308291386 |
